# COGNITIVE ASSESSMENT WITH COGNIVUE *CLARITY*^®^: PSYCHOMETRIC PROPERTIES AND ENHANCED NORMATIVE RANGES IN A DIVERSE POPULATION

**DOI:** 10.1101/2024.03.18.24304463

**Authors:** James E. Galvin, Lun-Ching Chang, Paul Estes, Heather M. Harris, Ernest Fung

## Abstract

**Background:** Detecting cognitive impairment in clinical practice is challenging as most instruments do not perform well in diverse samples of older adults. These same instruments are often used for eligibility into clinical trials making it difficult to recruit minoritized adults into Alzheimer’s disease (AD) studies. Cognivue *Clarity*^®^ is an FDA-cleared computerized cognitive screening platform using adaptive psychophysics to detect cognitive impairment.

**Objective:** Test the ability of Cognivue *Clarity* to detect cognitive impairment in a diverse community sample compared with the Repeatable Battery for the Assessment of Neuropsychological Status (RBANS).

**Methods:** This study enrolled 452 participants across 6 US study sites and completed both Cognivue *Clarity* device and RBANS. Psychometric properties and exploratory factor analysis of Cognivue *Clarity* were explored and comparisons against RBANS across different age, sex, education, and ethnoracial groups were conducted.

**Results:** Participants had a mean age of 47.9±16.1 years (range: 18-85), 63.6% were female, 45.9% had ≤12 years of education, 31.2% were African American and 9.2% were Hispanic. Cognivue *Clarity* had strong internal consistency, test-retest and minimal practice effects. A 4-factor structure (Memory, Attention, Visuomotor, and Discrimination) had excellent goodness of fit. Normalizing age effects improved performance. Race and education effects were similar to those seen with RBANS. Cognivue *Clarity* had strong correlation with RBANS.

**Conclusions:** Our study supports the use of Cognivue *Clarity* as an easy-to-use, brief, and valid cognitive assessment that can be used for identifying individuals with likely cognitive impairment in the clinical setting and those who could be candidates for AD research studies.

## INTRODUCTION

Alzheimer’s Disease and related dementias (ADRD) affect over 6.7 million people in the US [1] and more than 55 million people worldwide [2]. Detection of the early stages of ADRD is a clinical challenge with many people coming to medical attention and diagnosis at the moderate stage. Mild Cognitive Impairment (MCI) represents a prodromal state of ADRD, and recent reports suggest that nearly 80% of people living with MCI are never diagnosed [3]. The delays in diagnosis may be particularly relevant to individuals from racial and ethnic minorities, socioeconomically disadvantaged populations, and geographic locales that lack specialty memory care settings (i.e., rural areas) and who are at higher risk for ADRD [1]. There are likely many causes contributing to low rates of diagnosis including access to care, access to specialists, under- or non-insurance, disease stigma, and health literacy [4]. One potentially addressable cause is low rates of screening for cognitive impairment with a culturally valid and sensitive measure that does not require extensive staff or physician time.

Dementia screening could capture cases at the earliest possible stage in at-risk populations permitting early intervention and enrollment into trials [5,6]. However, at the present time, dementia screening is not endorsed by the US Preventive Services Task Force [7]. Reasons for this lack of recommendation include both lack of clear understanding of benefits vs harms of screening, and lack of agreement about the utility of commonly used screening tests such as the Mini Mental State Examination (MMSE) [8]. Although the Medicare Annual Wellness Visit (AWV) includes assessing for cognitive impairment as part of the required elements, there is no agreement on how this could be done and less than 25% of Medicare beneficiaries receive AWV as part of the medical care [9,10]. The most commonly used screening instruments used in the clinic and for clinical trial inclusion are the Mini Mental State Exam (MMSE) [8] and the Montreal Cognitive Assessment (MoCA) [11]. The MMSE has well recognized education and ethnoracial biases [12] including the serial subtraction task and lacks sensitivity to detecting MCI [13]. The MoCA also has educational and cultural biases including clock drawing, animal naming, and serial subtraction [14], which is only partly corrected by adding one point to those individuals with 12 years or less of education. Although a basic form of the MoCA was developed for low literacy populations [15], this format is likely not ideal for clinical trial screening.

The lack of screening and early detection, and delay in diagnosis of MCI and mild AD limits access of eligible older adults for treatment with newer disease modifying medications. Recently approved amyloid lowering therapies (i.e., Lecanemab) are only indicated for early-stage AD [16] and with most new cases diagnosed at the moderate stage [1,4], this represents a missed window of opportunity. Further, late detection limits the opportunity for interested and otherwise eligible older adults to participate in clinical trials to test new diagnostics and therapeutics, or in other clinical research projects to further our understanding of ADRD. Lastly, even when eligible individuals are identified and enrolled into clinical trials, the research sample rarely reflects the diversity of the US population. While racial and ethnic minorities comprise 39% of the US population [17], they account for only 2-16% of clinical trial participants [5, 18].

Of 4,105 US clinical trials registered in ClinicalTrials.gov from March 2000 to March 2020 that reported race/ethnicity data, the majority of enrollees were White (median 79.7%) [18], outpacing the White population in the 2020 US Census of 71% [17]. This same report found median participation in clinical trials at 10% for African Americans, 6% for Asians, and 0% for American Indians [18]. In a separate analysis [19], participation in one company’s clinical trials by Hispanic/Latino enrollees was 15.9%, though they represent 18.7% of the US population. Racial and ethnic minorities and socioeconomically disadvantaged groups have a higher lifetime risk of AD, experience a younger age of onset, present to medical attention at more advanced stages, and live more years with cognitive impairment [20]. Similar disparities are seen in between sex and educational level of the US population and that enrolled in clinical trials. Women make up 50.4% of the US population, but only 41.2% of participants in clinical trials [21]. Although 63% of US residents have less than a bachelor’s degree, 55.8% of participants in clinical trials had a college or advanced degree [22,23].

To address these unmet needs, we conducted a study of Cognivue *Clarity*^®^, the first FDA-cleared computerized cognitive test [24-26] that uses adaptive psychophysics to detect risk of cognitive impairment, in a diverse US sample across different age, education, sex, race and ethnicity strata. The Cognitive Testing in Diverse Populations to **<u>F</u>**urther the **<u>O</u>**bjective and **<u>C</u>**linical **<u>U</u>**nderstanding of Cognivue **<u>S</u>**tudy (FOCUS) study was used to calculate age-normed scores, assess performance across different sociodemographic variables, measure test-retest reliability and potential practice effects, and establish validity against a Gold Standard neuropsychological battery, the Repeatable Battery for the Assessment of Neuropsychological Status (RBANS), commonly used both in clinical practice and in clinical trials [27].

## METHODS

### Study Design

FOCUS was an open-label, single visit, multi-site validity and reliability study that enrolled participants to assess the psychometric properties and performance of Cognivue *Clarity* across a diverse population. Inclusion criteria were (a) age ≥18 years, (b) fluency in English, (c) community dwelling, (d) overall good health with no acute medical illness, (e) full vision in at least one eye, (f) full use of at least one hand, and (g) ability to provide informed consent. Exclusion criteria were: (a) presence of an acute medical illness, (b) residing in dependent care facility or in hospice, and (c) inability to be tested in English. Participants underwent a one-time study visit that lasted up to two hours. Following informed consent, each participant completed a sociodemographic questionnaire and was then randomized for assessment groups for 90-minutes of testing on 2 devices: Cognivue *Clarity* (10-minute assessment) or an Apple iPad to complete the RBANS (30-minute assessment) - either Cognivue *Clarity* followed by RBANS or RBANS followed by Cognivue *Clarity*. Each participant completed three consecutive trials of Cognivue *Clarity* with up to 5 minutes between each trial. The FOCUS study was deemed exempt by the Advarra Institutional Review Board (Pro00064617).

The primary objective of FOCUS was confirmation of scoring and normative ranges with the Cognivue *Clarity* across different ages, sexes, race and ethnic groups, and levels of education. Secondary study objectives included determination of the level of practice effects and psychometric properties, assessment of the overall score recommendations of Cognivue *Clarity*; and construct validity against a Gold Standard, the RBANS.

### Demographics and Self-Reported History

Participants provided self-reported information on age, sex, educational attainment (less than high school, high school degree or equivalent, associate degree, bachelor degrees, master or other graduate degree), racial identity (White, Black/African American, Asian, American Indian/Alaska Native, Native Hawaiian/Pacific Islander, Two or more races), ethnicity (Hispanic, Non-Hispanic), marital status, employment status, smoking history, alcohol consumption, social activity, exercise, hearing loss/use of hearing device, use of vision correction, and self-reported medical history (e.g., diabetes, hypertension, obesity, head injury, attention deficit disorder, cancer, COVID-19).

### Cognivue Clarity^®^

The automated Cognivue technology utilizes adaptive psychophysics assessing baseline motor skills and visual acuity to test information processing and eliminating biases that can be found in common cognitive testing mechanisms. Cognivue *Clarity* is comprised of 10 subtests consisting of two validity performance measures (Adaptive Motor Control, Visual Salience), four discrimination tests (Letter, Word, Shape, Motion), and four memory tests (Letter, Word, Shape, Motion). The Adaptive Motor Control subtest measures the speed at which the participant is able to manipulate the flywheel on the device. The Visual Salience subtest measures the threshold of the participant to determine visual contrast of the target stimuli. The Discrimination tests present a Letter, Word, Shape or Motion stimuli that the participant must match compared with non-target stimuli manipulating the flywheel. The Memory tests present a Letter, Word, Shape, or Motion stimuli the participant must first immediately recall and then tests delayed recall using a series of n-back paradigms. After completion, the Cognivue *Clarity* device provides immediate results in a report and/or in a comma-separated values file (**Figure 1**). Results include an overall average score, four domain specific scores, reaction times, and 10 subtest scores, as well as response tracings to assess performance validity. Score interpretations are provided as normal, borderline, or impaired performance.

**Figure 1:**
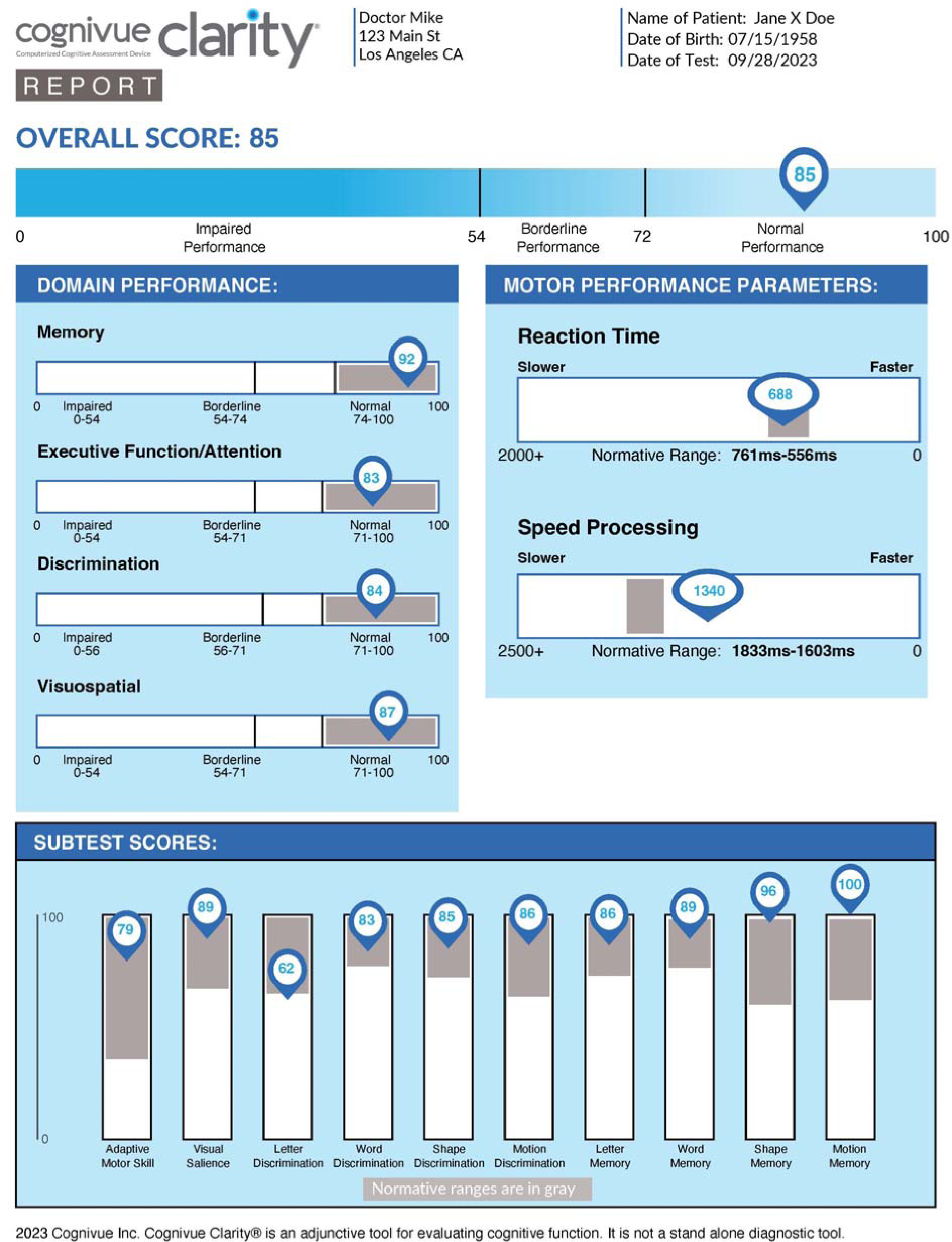
A sample report generated by Cognivue *Clarity*.

### Repeatable Battery for the Assessment of Neuropsychological Status (RBANS)

The RBANS (Pearson Assessments) was administered to each participant either before or after the Cognivue *Clarity*. The RBANS is commonly used as an assessment for dementia and other neurocognitive disorders and has been used both for screening and as an outcome in ADRD clinical trials and therefore was selected as Gold Standard for comparison. Four parallel forms are available with age-normed values from 12-89 years. The RBANS provides a total score, 5 index standard scores (immediate and delayed memory, visuospatial/construction, language, and attention), and can be completed in 30 minutes. An RBANS score of less than 85 is considered abnormal [27].

### Statistical Analyses

Statistical analyses were conducted using IBM SPSS v28 (Armonk, NY) and R (version 4.3.1). Descriptive statistics were used to summarize overall sample characteristics. Student t-tests or One-way analysis of variance (ANOVA) with Tukey-Kramer post-hoc tests were used for continuous data and Chi-square analyses were used for categorical data.

A total of 629 individuals completed the Cognivue *Clarity* component with 452 used in the analyses presented here. One individual did not have age recorded, 49 individuals had an undetermined educational attainment, and 127 individuals had questionable performance validity with score swings greater than 15 points between the three Cognivue *Clarity* trials. This is approximately 10% higher than the base rate of noncredible performance in clinical non-forensic settings [28-30]. Among the 452 participants, 110 had missing RBANS data, so the sample size for direct comparisons between Cognivue *Clarity* and RBANS was reduced to 342. A sensitivity analysis was performed removing 92 individuals who self-reported a neurological (e.g., head injury, transient ischemic attack, multiple sclerosis) or psychiatric (e.g., attention deficit disorder, depression, bipolar disorder) diagnosis. This was to ensure that Cognivue *Clarity* scores were not unduly influenced by neuropsychiatric conditions and to ensure results reflected a normative sample.

Internal consistency was examined as the proportion of the variability in the responses that is the result of differences in the respondents, reported as the Cronbach alpha reliability coefficient. Coefficients greater than 0.7 are good measures of internal consistency. Test-retest reliability was determined using the Spearman-Brown prediction formula.

A scree plot and exploratory factor analysis (EFA) with Oblique rotation was used to determine the individual factor loading of the 10 Cognivue subtests. Oblique rotation allows factors to be correlated with each other, frequently resulting in more interpretable factor loadings. We fit three possible models and examined the fit indices including Root Mean Square Error of Approximation with 90% confidence intervals (RMSEA), Chi-Square fit index, and the Tucker-Lewis Index (TLI). For RMSEA, values less than .05 are considered good. Ideally the Chi-Square test should be *non-*significant (with more than 0 degrees of freedom), however this is commonly not achieved in real data as sample sizes sufficient for factor analysis are also frequently large enough to detect even minor deviations between the estimated factor structure and the data. A good TLI is above 0.95. The factors derived from the EFA were then evaluated with a principal components analysis bi-plot which revealed 4 clear clusters replicating the EFA. A correlation heatmap was created to visualize associations between Cognivue Clarity average and subtest scores and RBANS total and index scores using Pearson product-moment correlation coefficients.

Group validity was assessed by examining Cognivue Clarity and RBANS by sample characteristics (sociodemographic and lifestyle characteristics, medical history). Linear regression was used to test the association between Cognivue *Clarity* and sociodemographic characteristics (e.g., age, race, education). To further study race effects, a stratified analyses was performed comparing performance on Cognivue Clarity and RBANS between White and Black participants. Unadjusted analyses demonstrated differences in age and education so adjusted analyses were performed. Cognivue Clarity was adjusted for age and education while RBANS, which is already age-normed, was adjusted for education only.

Cognivue Clarity scores were residualized on age to provide optimized age-norming for comparisons to RBANS scores which were already age-normed. Cognivue Clarity total and subtest scores were compared by cognitive status based on RBANS score cutoffs (<85) for cognitive impairment. Receiver operator characteristics (ROC) curves and area under the curve (AUC) were used to test discriminative properties of Cognivue and determine sensitivity, specificity, and optimal cut-points (by using closest top-left criteria). Multiple comparisons were addressed using the Bonferroni correction.

## RESULTS

### Sample Characteristics

FOCUS was conducted from September 8, 2022, to December 16, 2022, and included 452 subjects from 6 sites covering all 4 regions of the US (Northeast, Midwest, South, West) with valid Cognivue *Clarity* data. Participants had a mean age of 47.9±16.1 years (range: 18-85), 63.6% were female, 45.9% had 12 or less years of education, 35.9% were married, 41.9% were employed full time, 17.1% were current smokers, 20.0% consumed 2 or more alcohol drinks per week, 21.9% exercised daily, and 21.5% participated in daily social activities. The racial identity of the sample was 63.4% White, 31.2% Black/African American, while 5.4% reported other races. Hispanic ethnicity was reported in 9.2% of the sample. The sample self-reported common health problems including obesity (41.5%), diabetes (1.9%), hypertension (43.7%), cancer (4.8%), prior history of head injury (7.2%), vision correction (54.9%), hearing loss (3.7%), mood disturbance (6.3%), and COVID-19 (60.3%). The mean Cognivue *Clarity* score for the sample was 80.2±11.4 (range: 33-99) and the mean RBANS total score was 93.0±15.4 (range: 48-146)

### Psychometric Properties of Cognivue Clarity

Cognivue *Clarity* exhibited strong internal consistency and test-retest reliability. The Cronbach’s alpha was 0.812 (95%CI: 0.785-0.837); *p*<.001 and the test-retest reliability was 0.85. After eliminating individuals with exceptionally large swings between measurement occasions (subjects with between measurement differences of 15 or higher) the practice effect for Cognivue *Clarity* was 2.99 points per measurement, or approximately a 0.20 SD increase in score.

An exploratory factor analysis was conducted to establish the underlying data structure of Cognivue *Clarity*. The scree plot of the 10 subtests suggested a 1-, 3-, or 4-factor structure with fit indices shown in **Table 1**. The results suggest that not only is the 4-factor model the best fit, but it is also a *good* fit to the data, with all three fit indices in the acceptable range. The 4-factor model with factor loadings, sum of squares, eigenvalues, and variance are shown in **Table 2**. A delayed memory factor was the strongest, with high loadings from all memory scores. An executive attention factor (letter and word discrimination), a visuomotor factor (motor adaptive and visual salience) and a discrimination factor (shape and motion discrimination) were also defined. The factor structure had a cumulative variance of 0.52.

**Table 1:**
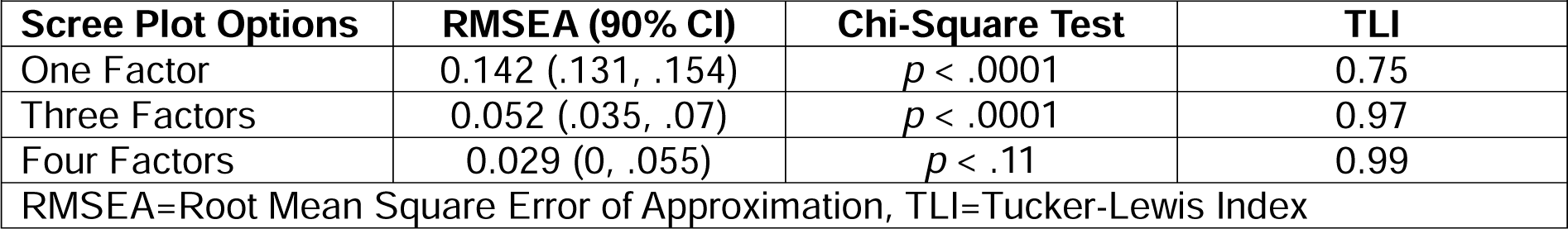
Goodness of Fit Indices.

**Table 2:** Factor Analysis of Cognivue *Clarity*.

| **Table 2: Factor Analysis of Cognivue *Clarity*** |  |  |  |  |
| --- | --- | --- | --- | --- |
| **Cognivue Subtests** | **Factor 1**<br>**Memory** | **Factor 2**<br>**Attention** | **Factor 3**<br>**Visuomotor** | **Factor 4**<br>**Discrimination** |
| Adaptive Motor |  |  | 0.81 |  |
| Visual Saliency |  |  | 0.61 |  |
| Letter Discrimination |  | 0.37 |  |  |
| Word Discrimination |  | 0.86 |  |  |
| Shape Discrimination |  |  |  | 0.39 |
| Motion Discrimination |  |  |  | 0.59 |
| Letter Memory | 0.67 |  |  |  |
| Word Memory | 0.85 |  |  |  |
| Shape Memory | 0.71 |  |  |  |
| Motion Memory | 0.48 |  |  |  |
| Eigenvalue | 3.89 | 1.37 | 0.99 | 0.75 |
| Sum of Squares Loading | 2.03 | 1.34 | 0.99 | 0.85 |
| Proportion Variance | 0.20 | 0.13 | 0.10 | 0.08 |
| Cumulative Variance | 0.20 | 0.34 | 0.44 | 0.52 |

### Cognivue Clarity Performance Across Sociodemographic Groups

**Table 3** shows the performance on Cognivue Clarity and RBANS by sociodemographic categories of age, education, sex, ethnicity, and race. There was no difference in Cognivue *Clarity* performance between men and women, or between individuals by ethnicity. There was a significant age effect (p<.001) with 18–39-year-olds performing best and individuals over age 70 performing worst. There were significant differences in performance by highest educational attainment (p<.001) with individuals with less than a Bachelor’s degree performing similarly and individuals with Bachelor’s degrees and graduate degrees performing similarly.

**Table 3:** Group Comparisons of Cognivue and RBANS Scores.

|  | Sex |  |  | Race |  |  | Ethnicity |  |  |
| --- | --- | --- | --- | --- | --- | --- | --- | --- | --- |
|  | Men | Women | p-value | White | Black | p-value | Non-Hispanic | Hispanic | p-value |
| Cognivue | 79.6 (12.0) | 80.6 (11.1) | .447 | 81.6 (10.7) | 77.0 (12.6) | <.001 | 80.2 (11.6) | 82.4 (9.4) | .143 |
| RBANS | 93.5 (16.6) | 92.8 (14.7) | .692 | 96.9 (14.3) | 85.2 (14.9) | <.001 | 93.4 (15.5) | 89.5 (14.3) | .208 |
|  | Age Categories |  |  |  |  |  |  |  |  |
|  | 18-29 | 30-39 | 40-49 | 50-59 | 60-69 | 70+ | p-value |  |  |
| Cognivue | 86.3 (8.8) | 85.3 (8.9) | 80.5 (11.6) | 79.9 (9.7) | 75.4 (11.8) | 70.8 (17.5) | <.001<sup>a</sup> |  |  |
| RBANS | 89.5 (15.6) | 86.2 (14.2) | 98.3 (13.6) | 94.5 (14.2) | 94.6 (15.9) | 93.1 (17.3) | <.001<sup>b</sup> |  |  |
|  | Highest Educational Attainment |  |  |  |  |  |  |  |  |
|  | <High School | High School | Associate | Bachelor | Graduate | p-value |  |  |  |
| Cognivue | 73.4 (14.1) | 79.2 (12.0) | 79.5 (11.6) | 83.6 (9.6) | 82.9 (8.1) | <.001<sup>c</sup> |  |  |  |
| RBANS | 83.8 (9.8) | 87.4 (15.3) | 94.0 (12.1) | 97.1 (12.7) | 106.2 (15.2) | <.001<sup>d</sup> |  |  |  |

Mean (SD), **Bold** p-values are significant after correction for multiple comparisons (adjusted p-value=0.01).

For race, only comparisons between White and Black participants were considered since there were too few participants in other racial groups to provide meaningful conclusions. There were significant differences between White and Black participants on Cognivue *Clarity* (p<.001). There was no interaction between race and education supporting that Cognivue *Clarity* performance differences in Black participants are not due to differences in education. Differences in scores were further analyzed by conducting stratified analyses comparing White and Black participants by Cognivue Clarity average score and subtests, and RBANS total score and index scores (**Table 4**). In unadjusted analyses for Cognivue *Clarity*, African American participants performed worse than White participants in the average score and the motion discrimination, shape memory, and motion memory subtests. After adjusting for age and education, adaptive motor scores were also different. For RBANS, the total score and all index scores were different in unadjusted analyses with only delayed memory index losing significance after adjusting for education.

**Table 4:** Race Stratified Analyses for Cognivue Clarity and RBANS.

| **Table 4: Race Stratified Analyses for Cognivue Clarity and RBANS** |  |  |  |  |
| --- | --- | --- | --- | --- |
| **Variable** | **White** | **Black** | **Unadjusted p-value** | **Adjusted p-value<sup>a</sup>** |
| Age | 50.9 (16.3) | 45.7 (15.6) | **.009** | ---- |
| Education, % 12y or less | 37.0 | 64.0 | **<.001** | ---- |
| Cognivue Average Score | 81.9 (10.7) | 76.4 (12.7) | **<.001** | **<.001** |
| Cognivue Adaptive Motor | 55.3 (15.6) | 49.6 (17.3) | .008 | **<.001** |
| Cognivue Visual Salience | 80.7 (13.1) | 79.8 (14.6) | .612 | .089 |
| Cognivue Letter Discrimination | 75.7 (14.0) | 72.0 (15.2) | .055 | .020 |
| Cognivue Word Discrimination | 78.3 (16.2) | 78.2 (15.4) | .967 | .330 |
| Cognivue Shape Discrimination | 86.9 (14.5) | 81.7 (16.8) | .012 | .004 |
| Cognivue Motion Discrimination | 83.8 (18.6) | 73.9 (21.7) | **<.001** | **<.001** |
| Cognivue Letter Memory | 82.6 (16.6) | 81.1 (16.6) | .460 | .407 |
| Cognivue Word Memory | 88.9 (15.8) | 83.7 (19.3) | .026 | .008 |
| Cognivue Shape Memory | 80.6 (21.4) | 71.4 (24.9) | **.002** | **.001** |
| Cognivue Motion Memory | 82.3 (22.5) | 71.8 (29.8) | **.003** | **.002** |
| RBANS Total Score | 97.3 (14.4) | 85.7 (15.1) | **<.001** | **<.001** |
| RBANS Immediate Memory Index | 91.6 (16.5) | 81.5 (16.8) | **<.001** | **<.001** |
| RBANS Delayed Memory Index | 97.8 (16.3) | 90.6 (18.0) | **.001** | .016 |
| RBANS Attention Index | 102.3 (17.9) | 93.9 (19.3) | **<.001** | **.005** |
| RBANS Visuospatial Index | 103.4 (15.5) | 91.6 (18.0) | **<.001** | **<.001** |
| RBANS Language Index | 95.1 (13.0) | 87.6 (16.0) | **<.001** | **.003** |
| Mean (SD) or %, **Bold** p-values are significant after correction for multiple comparisons (adjusted p=.0045 for Cognivue; adjusted p=.0083 for RBANS) |  |  |  |  |
| <sup>a</sup>Cognivue adjusted for age and education, RBANS adjusted for education only |  |  |  |  |
| RBANS = Repeatable Battery Assessing Neuropsychological Status |  |  |  |  |

### Age Adjustment of Cognivue Clarity

To explore the age effect, a simple linear regression was fitted and demonstrated a strong association between age and Cognivue *Clarity* performance. Increasing age decreases the average Cognivue *Clarity* score (94.38 – 0.297 x age, R^2^=17.2%). We also fit a multiple linear regression model with race and education and found that White participants scored on average 5.47 points higher than Black participants matched for age and educational attainment (R^2^ = 27.8%). Education did not provide additional fit to the regression model.

### Comparison of Cognivue Clarity Performance to RBANS

Since no formal clinical evaluation was included in FOCUS, the RBANS was treated as a gold standard for cognitive performance with a cut-point of 85 to represent possible cognitive impairment. A total of 342 participants had complete and valid Cognivue *Clarity* and RBANS data available. **Table 3** shows the RBANS performance across the different sociodemographic groups with significant differences by education and race, similar to what is seen with Cognivue *Clarity.* The Pearson’s correlation between Cognivue *Clarity* and RBANS score was 0.385 with an AUC 0.666 (95%CI: 0.601 – 0.731) for RBANS impaired versus RBANS not impaired. Since RBANS scores are provided as age-normed scores, residualizing the Cognivue *Clarity* score on age improved the correlation to 0.495, and the AUC to 0.730 AUC. To achieve the optimal cut point age-normed Cognivue *Clarity* score (residual) based on the post-hoc analyses from **Table 1**, we categorized age into three groups (18 – 39, 40 – 59 and 60+), then de-age normed back the original Cognivue *Clarity* score. **Table 5** shows the AUC, sensitivity, specificity, and optimal cut for Cognivue *Clarity* score for each age group.

**Table 5:** Age-Strata Performance of Cognivue Compared with RBANS.

| Table 5: Age-Strata Performance of Cognivue Compared with RBANS |  |  |  |  |  |  |  |  |  |
| --- | --- | --- | --- | --- | --- | --- | --- | --- | --- |
|  | Age 18-39 |  |  | Age 40-59 |  |  | Age 60+ |  |  |
|  | RBANS<br>Not Impaired | RBANS<br>Impaired | p-value | RBANS<br>Not Impaired | RBANS<br>Impaired | p-value | RBANS<br>Not Impaired | RBANS<br>Impaired | p-value |
| Cognivue Average Score | 88.8 (5.8) | 82.4 (9.2) | **<.001** | 82.5 (9.2) | 74.1 (10.8) | **<.001** | 76.9 (9.7) | 64.9 (13.8) | **<.001** |
| RBANS Total Score | 98.1 (8.7) | 74.1 (9.7) | **<.001** | 101.3 (9.6) | 75.3 (9.5) | **<.001** | 101.1 (13.3) | 76.3 (6.4) | **<.001** |
| Impaired by Cognivue, % | 19.4 | 53.3 | **<.001** | 34.9 | 76.0 | **<.001** | 39.3 | 60.7 | **<.001** |
| Area Under the Curve (95% CI) | .695 (0.593-0.797) |  |  | 0.716 (0.613-0.819) |  |  | 0.750 (0.651-0.849) |  |  |
| Best Cut Point | 86 |  |  | 81 |  |  | 68 |  |  |
| Sensitivity | 65.1 |  |  | 65.8 |  |  | 88.4 |  |  |
| Specificity | 65.1 |  |  | 72.4 |  |  | 57.1 |  |  |
| Mean (SD) or %, **Bold** p-values are significant after correction for multiple comparisons (adjusted p-value=0.017).<br>RBANS= Repeatable Battery Assessing Neuropsychological Status |  |  |  |  |  |  |  |  |  |

We conducted a Principal Component Analysis (PCA) bi-plot of age-normed scores from the 10 Cognivue *Clarity* subtests with RBANS scores and categorized participants into four clear subgroups consistent with domains derived from the factor analysis (data not shown). We then calculated the correlation of the 10 age-normed subtests scores with the five RBANS indexes (immediate memory, delayed memory, visuospatial, attention and language). A correlation heatmap was created to visualize associations between Cognivue *Clarity* average and subtest scores and to RBANS total and index scores using Pearson product-moment correlation coefficients. **Figure 2** shows that the motion memory, shape memory and word memory scores are highly correlated with all RBANS indexes except language index, while the letter memory score is more weakly correlated with these same RBANS indexes. The motor, visual, motion discrimination and shape discrimination scores are correlated with RBANS visuospatial/constructional index and attention domain index, while the letter discrimination and word discrimination scores are correlated with RBANS attention domain index only.

**Figure 2:**
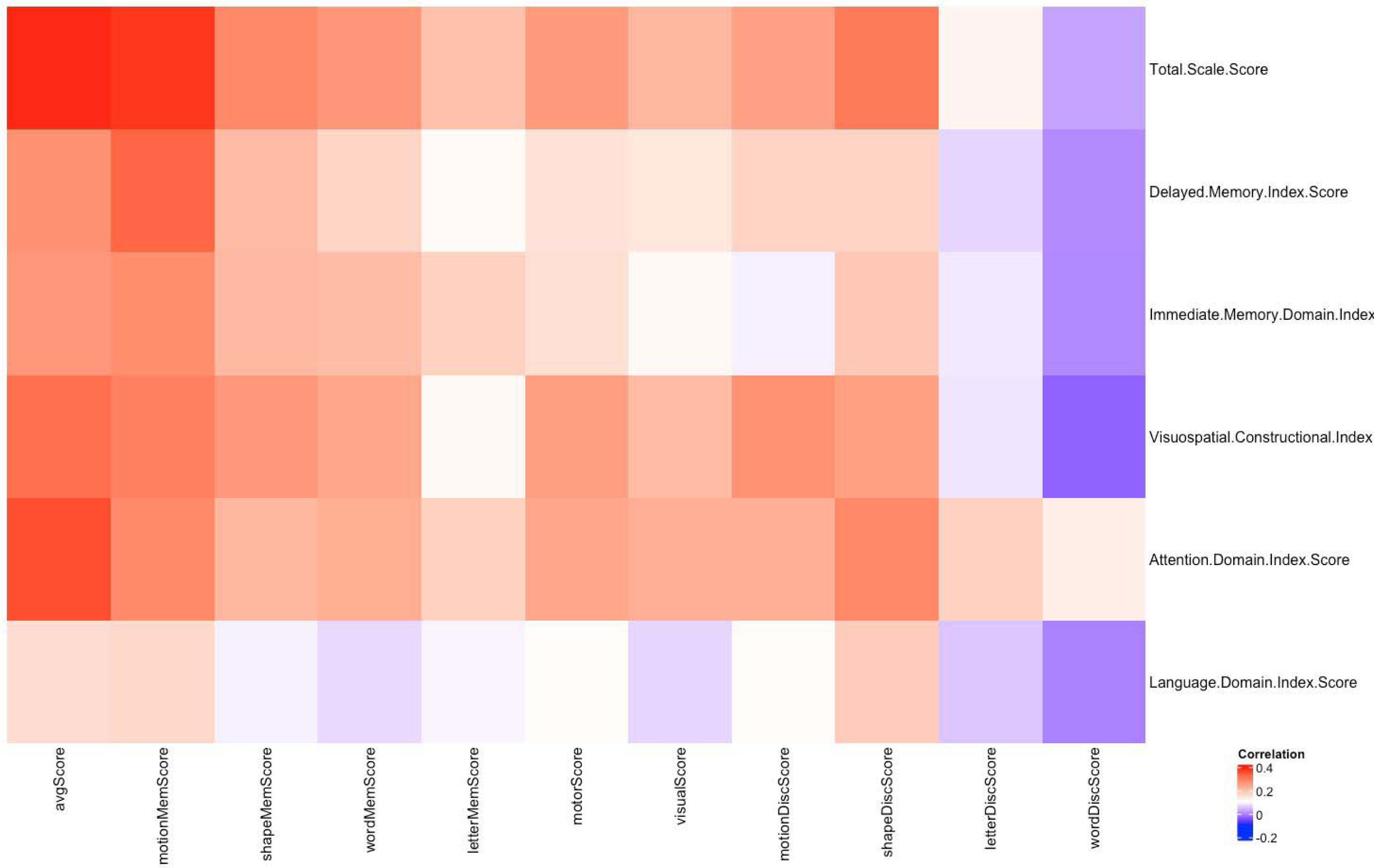
Correlation Map for Cognivue *Clarity* and RBANS. A correlation heatmap was created to visualize associations between Cognivue *Clarity* average and subtest scores and RBANS total and index scores using Pearson product-moment correlation coefficients. Motion memory, shape memory and word memory scores are highly correlated with all RBANS indexes except language index, while correlation is weaker between letter memory score and these same RBANS indexes. The motor, visual, motion discrimination and shape discrimination scores are correlated with RBANS visuospatial/constructional index and attention domain index, while the letter discrimination and word discrimination scores are correlated with RBANS attention domain index only.

### Sensitivity Analysis

While only self-reported medical history was available, 92 individuals reported neurologic (e.g., head injury, stroke) or psychiatric (e.g., attention deficit disorder, bipolar disorder, depression) conditions. These individuals had a mean age 46.9±15.7 (range 18-73), 23.9% were African American, 11.9% were Hispanic, and 41.3% had 12 years or less of education. To ensure that our results were not influenced by the performance of these 92 individuals, we repeated the analyses comparing Cognivue *Clarity* scores by RBANS status after removing the 92 individuals. As shown in **Table 6**, there was no difference in Cognivue *Clarity* performance on average scores or 10 subtest scores between the original and sensitivity samples. The RBANS total score and age were also similar between the two samples.

**Table 6:** Sensitivity Analysis of Cognivue *Clarity*.

| Variable | Original Sample (n=452) |  |  | Sensitivity Sample (n=360) |  |  |
| --- | --- | --- | --- | --- | --- | --- |
|  | RBANS<br>Not Impaired | RBANS<br>Impaired | p-value | RBANS<br>Not Impaired | RBANS<br>Impaired | p-value |
| Age | 49.5 (15.9) | 45.6 (17.3) | .044 | 50.1 (15.9) | 46.1 (17.6) | .08 |
| Cognivue Average Score | 82.4 (9.7) | 75.2 (13.3) | **<.001** | 82.3 (9.9) | 74.9 (13.6) | **<.001** |
| Cognivue Adaptive Motor | 56.3 (16.1) | 49.2 (16.7) | **<.001** | 55.5 (15.9) | 48.7 (16.3) | **.001** |
| Cognivue Visual Saliency | 82.2 (12.6) | 77.8 (14.1) | **.005** | 81.7 (13.0) | 77.8 (14.6) | **.030** |
| Cognivue Letter Discrimination | 75.8 (13.3) | 72.8 (16.3) | .083 | 75.6 (13.4) | 72.1 (16.9) | .091 |
| Cognivue Word Discrimination | 78.0 (15.1) | 78.8 (16.7) | .653 | 78.4 (15.4) | 78.4 (17.3) | .980 |
| Cognivue Shape Discrimination | 87.8 (13.1) | 79.1 (18.2) | **<.001** | 87.7 (13.3) | 79.2 (18.5) | **<.001** |
| Cognivue Motion Discrimination | 83.2 (19.2) | 76.1 (21.1) | **.003** | 82.9 (19.3) | 75.1 (21.4) | .004 |
| Cognivue Letter Memory | 83.4 (15.8) | 79.6 (17.5) | .079 | 83.1 (15.8) | 79.5 (17.4) | .108 |
| Cognivue Word Memory | 89.4 (14.5) | 80.3 (20.5) | **<.001** | 89.4 (14.8) | 81.0 (20.3) | **<.001** |
| Cognivue Shape Memory | 81.0 (19.8) | 69.7 (27.2) | **<.001** | 80.9 (19.9) | 69.3 (27.7) | **<.001** |
| Cognivue Motion Memory | 84.5 (21.3) | 68.4 (29.2) | **<.001** | 83.9 (21.7) | 67.6 (30.0) | **<.001** |
| RBANS Total Score | 100.4 (10.7) | 75.0 (8.8) | **<.001** | 100.8 (10.6) | 75.1 (8.6) | **<.001** |
| Mean (SD), **Bold** p-values are significant after correction for multiple comparisons (adjusted p=.0036)<br>RBANS = Repeatable Battery Assessing Neuropsychological Status |  |  |  |  |  |  |

## DISCUSSION

Cognivue *Clarity* offers a rapid, valid, and reliable measure of cognitive function with limited practice effects due to prior testing in a study of community-dwelling individuals from age 18-85. This suggests that Cognivue *Clarity* can be used to screen individuals for cognitive impairment and follow cognitive performance over time. Cognivue *Clarity* has a 4-factor structure that provides not only a global performance score but also information on delayed memory, executive-attention, visuomotor, and discrimination-perceptual abilities. Cognivue *Clarity* performed equally well between men and women but had age effects with improved discriminability following age-norming. African Americans scored 5.5 points lower than White participants matched for age and education. At least some of these differences can be explained by differential performance in motion discrimination, motion memory, and shape memory subtests but further research is warranted.

Cognivue *Clarity* average score and subtests showed good correlation with RBANS total score and 4 of 5 index scores consistent with the 4-factor structure. RBANS Language index scores showed weak to no correlation with Cognivue *Clarity* average score or subtests scores. Using a cut-off of 85 on the RBANS total score, we were able to explore sensitivity and specificity of Cognivue *Clarity*. However, we do not have sufficient information on the sample to explain reasons for impaired performance on the Cognivue *Clarity* or RBANS in individuals aged 18-59. Further studies of individuals with established diagnoses will be required to address this. However, Cognivue *Clarity* performed similarly to RBANS across different age, education, sex, race, and ethnicity groups.

The detection of MCI and AD is limited in community settings [31-33] due in part to the use of brief screening tests of objective cognitive performance that lack psychometric properties similar to those of Gold Standard tests [4,34]. However, Gold Standard tests, such as the RBANS, are lengthy and require specialized staff to administer, score and interpret, making them impractical in the clinic. Cognivue *Clarity* offers a global assessment of cognitive functioning for easy screening while also providing domain and subtest scores and reaction times for more detailed characterization of patients. Cognivue *Clarity* adaptive psychophysics also provides the potential for use in clinical research with good test-retest reliability, internal consistency, and small practice effects for repeated testing in the context of a clinical trial [35].

The FOCUS study, which recruited a patient population that was diverse in age, race, and level of educational attainment, enhances our understanding regarding the normative ranges in cognitive assessment. This is important as the proportion of US adults who are diverse is increasing [36]. Historically underserved and underrepresented groups constitute 39% of the US population and FOCUS was able to closely match these demographic characteristics as well as educational attainment in which 63% of the US population has less than a Bachelor’s degree [37]. Studies that include diverse cohorts will enable the development of more diverse normative ranges and more precise interpretations of cognitive tests. This can further inform the use of dementia screening tools such as the Cognivue *Clarity* in clinical practice for early detection and in research for clinical trial eligibility.

### Study Limitations

FOCUS was a cross-sectional study so that longitudinal changes in Cognivue *Clarity* were not able to be discerned. Health history was self-reported, so no independent verification was available. A greater than expected number of individuals under age 60 had difficulty with both the Cognivue *Clarity* and RBANS, but in the absence of a comprehensive clinical evaluation the reasons remain unclear. Future research projects will be needed to test Cognivue *Clarity* properties in diagnosed individuals. One such study is the Bio-Hermes study conducted by the Global Alzheimer’s Platform [26] that includes characterized individuals with digital, imaging and plasma AD biomarkers, including Cognivue *Clarity*. While FOCUS had good representation of Non-Hispanic White, African American and Hispanic individuals from age 18-85, no conclusions about Cognivue *Clarity* performance can be determined regarding other races and ethnicities. Future research in these understudied groups is needed with several studies already underway.

## Conclusions

FOCUS was able to recruit a diverse sample that reflects many of the demographic characteristics of the US population. The psychometric properties of Cognivue *Clarity* suggests that it could play a significant role in early detection and screening for cognitive impairment across diverse populations capturing both global performance as well as domain- and subtest-specific attributes. This study supports the use of Cognivue *Clarity* as an easy-to-use, brief, and valid cognitive assessment that can be used for identifying individuals with likely cognitive impairments and individuals who could be candidates for clinical research studies.

## AUTHOR CONTRIBUTIONS

**Dr. Galvin** was involved in the formal analysis, methodology, writing of original draft, and review and editing the final manuscript. He approves of the final version and ensures the accuracy and integrity of the work.

**Dr. Chang** was involved in the formal analyses, writing of original draft, and review and editing the final manuscript. He approves of the final version and ensures the accuracy and integrity of the work.

**Mr. Estes** was involved in the conceptualization, methodology, and review and editing the final manuscript. He approves of the final version and ensures the accuracy and integrity of the work.

**Ms. Harris** was involved in the conceptualization, methodology, and review and editing the final manuscript. She approves of the final version and ensures the accuracy and integrity of the work.

**Dr. Fung** was involved in the review and editing the final manuscript. He approves of the final version and ensures the accuracy and integrity of the work.

## ACKNOWLEDGEMENTS AND FUNDING SOURCES

This study was funded by Cognivue, Inc. The authors acknowledge the research support provided by Velocity Clinical Research and thank the following 14 Velocity Clinical Research Sites for their precision and timely execution: Anderson-SC, Austin-TX, Boise-ID, Cincinnati-OH, Denver-CO, Gaffney-SC, Grants Pass-OR, New Smyrna Beach-FL, North Hollywood-CA, Providence-RI, San Diego-CA, Spartanburg-SC, Spokane-WA, Syracuse-NY. In addition, we acknowledge and thank the following Velocity Clinical Research partners for their contributions to ensuring timely, precise, and diverse patient participation: Charles-Herbert Devaux, Emily Kelly, Jennifer Carl and Erin Williams. We also acknowledge the following Cognivue Working Group Members for their contributions to the development, implementation, and analysis of the FOCUS study: Catherine Tallmadge, Shiva Pal, Seth Wideman, Jennifer Stubbs, Rob Parody, PhD, and Joel Raskin, MD.

## DATA AVAILABILITY STATEMENT

The dataset for this project is available to all interested parties. Please contact JEG at.

